# Leveraging Deep Learning of Chest Radiograph Images to Identify Individuals at High Risk for Chronic Obstructive Pulmonary Disease

**DOI:** 10.1101/2024.11.14.24317055

**Authors:** Saman Doroodgar Jorshery, Jay Chandra, Anika S. Walia, Audra Stumiolo, Kristin Corey, Seyedeh Maryam Zekavat, Aniket N. Zinzuwadia, Krisha Patel;, Sarah Short, Jessica L. Mega, R. Scooter Plowman, Neha Pagidipati, Shannon S. Sullivan, Kenneth W. Mahaffey, Svati H. Shah, Adrian F. Hernandez, David Christiani, Hugo J.W.L. Aerts, Jakob Weiss, Michael T. Lu, Vineet K. Raghu, the Project Baseline Health Study Group

## Abstract

**Background:** This study assessed whether deep learning applied to routine outpatient chest X-rays (CXRs) can identify individuals at high risk for incident chronic obstructive pulmonary disease (COPD).

**Methods:** Using cancer screening trial data, we previously developed a convolutional neural network (CXR-Lung-Risk) to predict lung-related mortality from a CXR image. In this study, we externally validated CXR-Lung-Risk to predict incident COPD from routine CXRs. We identified outpatients without lung cancer, COPD, or emphysema who had a CXR taken from 2013-2014 at a Mass General Brigham site in Boston, Massachusetts. The primary outcome was 6-year incident COPD. Discrimination was assessed using AUC compared to the TargetCOPD clinical risk score. All analyses were stratified by smoking status. A secondary analysis was conducted in the Project Baseline Health Study (PBHS) to test associations between CXR-Lung-Risk with pulmonary function and protein abundance.

**Findings:** The primary analysis consisted of 12,550 ever-smokers (mean age 62·4±6·8 years, 48.9% male, 12.4% rate of 6-year COPD) and 15,298 never-smokers (mean age 63·0±8·1 years, 42.8% male, 3.8% rate of 6-year COPD). CXR-Lung-Risk had additive predictive value beyond the TargetCOPD score for 6-year incident COPD in both ever-smokers (CXR-Lung-Risk + TargetCOPD AUC: 0·73 [95% CI: 0·72-0·74] vs. TargetCOPD alone AUC: 0·66 [0·65-0·68], p<0·01) and never-smokers (CXR-Lung-Risk + TargetCOPD AUC: 0·70 [0·67-0·72] vs. TargetCOPD AUC: 0·60 [0·57-0·62], p<0·01). In secondary analyses of 2,097 individuals in the PBHS, CXR-Lung-Risk was associated with worse pulmonary function and with abundance of SCGB3A2 (secretoglobin family 3A member 2) and LYZ (lysozyme), proteins involved in pulmonary physiology.

**Interpretation:** In external validation, a deep learning model applied to a routine CXR image identified individuals at high risk for incident COPD, beyond known risk factors.

**Funding:** The Project Baseline Health Study and this analysis were funded by Verily Life Sciences, San Francisco, California.

**ClinicalTrials.gov Identifier:** NCT03154346

## INTRODUCTION

Chronic obstructive pulmonary disease (COPD) is characterized by persistent respiratory symptoms (shortness of breath, chronic cough, phlegm production) due to airway and/or alveoli abnormalities that cause prolonged, progressive airflow obstruction.^1^ COPD is the third leading cause of mortality worldwide^2^ and carries an estimated burden of $30 billion annually to the United States (U.S.) healthcare system.^3^ COPD is incurable; however, early diagnosis and subsequent lifestyle^4^ and pharmaceutical intervention^5^ improve prognosis.^6^ The key diagnostic criterion for COPD is a post-bronchodilator forced expiratory volume in one second (FEV_1_) to forced vital capacity (FVC) ratio ≤0·7 as measured by spirometry.^7^

Current guidelines do not recommend screening asymptomatic adults for COPD.^7,8^ Instead, targeted case finding using spirometry has been proposed for individuals with a high suspicion of COPD: patients with chronic respiratory symptoms, structural abnormalities of the airways, and prevalent risk factors (e.g., smoking, exposure to pollutants).^7^ However, spirometry is often unavailable in low-income countries^9,10^ and is underutilized with disparate accessibility in high-income countries,^11^ leading to estimates that 50%-75% of COPD cases remain undiagnosed.^12,13^ Although patients with undiagnosed COPD are typically at an earlier disease stage,^14^ these individuals have a similar risk of mortality to those with confirmed COPD.^12^ Identifying undiagnosed cases could help preserve quality of life in these patients by enabling targeted interventions to slow disease progression.^8^

Several approaches have been proposed to identify patients at high risk for COPD.^15^ Most focus on surveys or questionnaires administered to patients during routine primary care visits.^16,17^ Another promising approach is to opportunistically identify high-risk individuals using routinely collected data in the electronic medical record (EMR), such as demographics, history of respiratory disease, and smoking history.^15,18,19^ Smoking is a major driver of COPD risk but is often not documented or recorded inaccurately in the EMR,^20^ and a growing proportion of COPD cases occur in never-smokers, for whom there are fewer well-established risk factors.^21^

Chest radiographs (CXRs) are one of the most common diagnostic tests in medicine^22^ and are a first-line imaging test for respiratory symptoms, including in primary and urgent care settings. Recent advances in artificial intelligence (AI), especially convolutional neural networks (CNNs),^23^ have enabled breakthroughs in extracting information from a CXR image to assess disease risk.^24,25^ We previously developed an AI model called CXR-Lung-Risk that can estimate the risk of 18-year lung-related mortality (COPD, lung cancer, interstitial lung disease, chronic emphysema) based on a single posterior-anterior (PA) CXR image as the only input.^26^ This model was validated in multiple clinical trial cohorts, and the CXR-Lung-Risk output was associated with survival in lung cancer patients.

Here, we tested whether the CXR-Lung-Risk model can be applied to routine, outpatient CXR images to identify patients in the EMR at high risk for incident COPD (Fig. 1). We compared the performance of the CXR-Lung-Risk model with an EMR-based clinical risk score (TargetCOPD).^19^ Additionally, we leveraged CXR images from participants in the Project Baseline Health Study (PBHS) to test whether the CXR-Lung-Risk score was associated with lower lung function and plasma protein concentrations. CXR is a widely used imaging modality; therefore, these findings may show the potential for opportunistic screening using deep learning– based models to identify high-risk individuals and guide COPD prevention.

**Figure 1.**
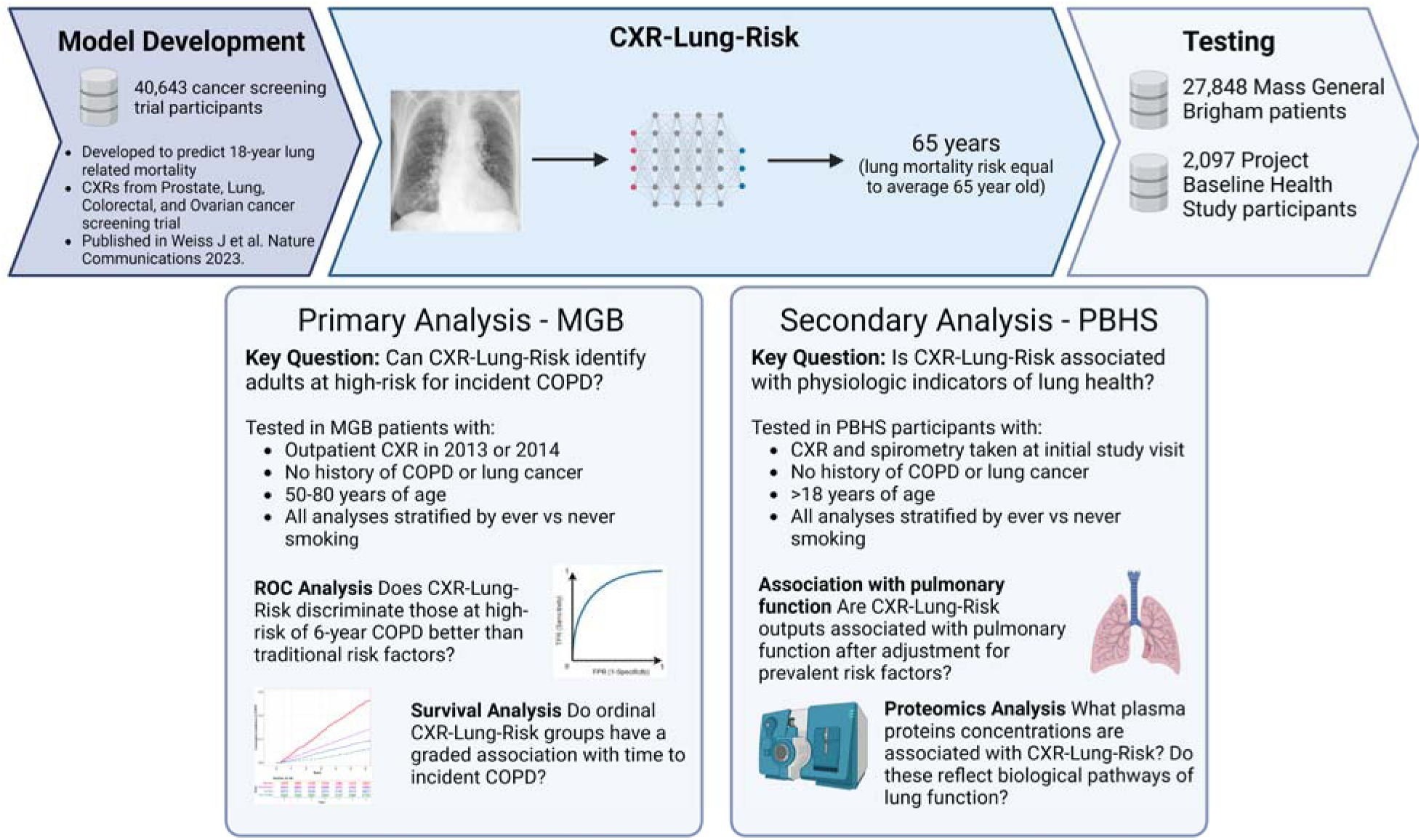
Schematic diagram of the study.

## METHODS

### Cohort description, sample inclusion, and exclusion criteria

Our primary analysis included 27,848 outpatients (Supplementary Fig. 1) ages 50–80 who had a posterior-anterior (PA) CXR taken at a Massachusetts General Brigham (MGB) hospital from 2013–2014 and no history of lung cancer, COPD, or chronic emphysema as defined by the International Classification of Diseases, 9th and 10th revision (ICD-9 and ICD-10) diagnosis codes (Supplementary Table 1). Analyses were performed in sub-cohorts stratified by ever-smokers (N=12,550) vs. never-smokers (N=15,298). This study was approved by the Mass General Brigham Institutional Review Board with a waiver of informed consent for retrospective analysis of deidentified data. The study followed Transparent Reporting of a Multivariable Prediction Model for Individual Prognosis or Diagnosis (TRIPOD) reporting guidelines for a risk prediction model validation study.^42^

In secondary analyses, we tested the association of CXR-Lung-Risk with pulmonary function testing and plasma protein abundances using data from the Project Baseline Health Study (PBHS),^43^ a diverse cross-sectional cohort study across four U.S. sites (sponsored by Verily Life Sciences) (ClinicalTrials.gov Identifier: NCT03154346). The deeply phenotyped PBHS cohort consists of 2,502 participants enriched for lung cancer and cardiovascular disease risk factors with demographic, survey, clinical, molecular, laboratory, and imaging taken at the initial study visit. Of the 2,502 patients, 2,097 had PA CXR images available (Fig. 1), and 1,263 underwent pulmonary function testing. 957 participants had CXR imaging and proteomic data available for 289 plasma proteins. Individuals with known lung cancer or prevalent COPD were removed from all analyses.

### CXR-Lung-Risk and chest radiograph images

The CXR-Lung-Risk model was developed to predict a composite outcome of 18-year lung disease (COPD, lung cancer, interstitial lung disease, chronic emphysema) mortality based on a single CXR image using 147,497 images from the Prostate, Lung, Colorectal, and Ovarian (PLCO) Cancer Screening Trial.^26^ The output of the model is expressed in years rather than a percentage to enhance interpretability (e.g., a CXR-Lung-Risk score of 60 years means the individual has lung-related mortality risk equivalent to the average 60-year-old). The PLCO study was conducted between 1993 and 2001 at 10 U.S. sites. This model was validated in two held-out testing datasets not used during model development. These radiographs were obtained from asymptomatic volunteers for lung cancer screening trials. Here, we tested the CXR-Lung-Risk model in a patient cohort where radiographs were obtained during routine care.

This study serves as an external validation of CXR-Lung-Risk using the existing free, open-source version without alterations (https://github.com/AIM-Harvard/CXR-Lung-Risk). Routine CXR images from MGB patients were obtained from the Picture Archiving and Communication System (PACS). For patients with multiple radiographs, we used the earliest radiograph during the study period. We expected no meaningful overlap of PLCO participants with our analysis cohort since no PLCO study site was in the same region as our current analysis cohort, and the last PLCO participant was enrolled 12 years before the MGB cohort and 16 years before the PBHS study began.

### Smoking history, risk factors, race, and ethnicity

In the MGB cohort, information about smoking history and risk factors, including recent dyspnea and medication use, was collected from the EMR. Smoking status was determined through manual review of clinical history, physical exam, and pulmonary function reports. A previously described algorithm^44^ was used to extract pack-years at the time of the CXR image for each patient. Patients without any smoking information were considered never-smokers. The presence of COPD and other comorbidities were identified using ICD-9 and ICD-10 codes (Supplementary Table 1).^45,46^ Race and ethnicity information was based on self-reported data and followed the guidelines outlined in the National Institutes of Health Policy on Reporting Race and Ethnicity Data.^47^ In the PBHS cohort, all risk factors were based on self-reported data.

### Outcomes

The primary outcome was incident COPD in the 6 years after the initial chest radiograph based on combined ICD-9 and ICD-10 codes obtained from the EMR (Supplementary Table 1). All-cause mortality was determined using the Social Security Master Death Index and the Mass General Brigham death registration system.

### TargetCOPD clinical risk score

We compared CXR-Lung-Risk to the TargetCOPD score,^19^ a regression model that includes age, smoking status, dyspnea, prescriptions for short-acting beta agonist (SABA), and prescriptions for antibiotics to output a probability that an individual has COPD. The TargetCOPD score was developed and validated from a large cluster randomized controlled case finding trial in primary care to predict the risk of undiagnosed COPD using data from the EMR. For binary analyses, we used this score with the published binary threshold of ≥7·5% risk. We assessed whether CXR-Lung-Risk had added value to predict incident COPD beyond the TargetCOPD score. Patients without medication information were considered to have no prescriptions for salbutamol or antibiotics (0·7% of the cohort).

### Protein abundance data

Plasma proteins from 957 participant samples from the PBHS study were used to associate CXR-Lung-Risk with underlying biologic disease mechanisms. Each plasma protein sample from the PBHS study was prepared using microflow high-resolution liquid chromatography-mass spectrometry. Then, these raw data were converted to protein abundances through the use of Dia-NN, v1.8.1 (https://github.com/vdemichev/DiaNN). Detailed steps of the quality control process can be found in the Supplementary Material. A total of 289 proteins were detected across all patient plasma samples. Microbial proteins, contaminants, and Ig variable chain proteins were not included in the analyses.

### Statistical analyses

We assessed the discrimination of CXR-Lung-Risk vs. the TargetCOPD score using time-dependent area under the receiver operating characteristic curves (AUC) over 6-, 3-, and 1-year follow-up periods. We used DeLong’s method to calculate the confidence intervals (CIs) for all AUCs. To address censoring, we assessed the association of the CXR-Lung-Risk score with incident COPD using Cox proportional hazards survival analysis, adjusted for clinical variables including age, smoking status, recent dyspnea, SABA use, prevalent asthma, antibiotic use, and findings from the CXR report, including the presence of a lung opacity, atelectasis, pneumothorax, pneumonia, edema, consolidation, or a lung lesion. We stratified the continuous CXR-Lung-Risk score into three ordinal groups (low risk: ≤50; moderate risk: >50 to ≤55; and high risk: >55) in both ever- and never-smokers. Cumulative incidence curves were calculated to assess the association of the ordinal risk groups with COPD incidence. All analyses were stratified by smoking status (ever-vs. never-smokers).

Secondary analyses were conducted in PBHS participants with pulmonary function tests (PFTs) and plasma proteomics. We related CXR-Lung-Risk scores to percentage of predicted peak expiratory flow (PEF), percentage of predicted forced vital capacity (FVC), percentage of predicted FEV_1_, FEV_1_/FVC ratio and abundance of 289 plasma proteins. Linear regression was used to adjust for age, sex, self-reported race, respiratory disease, body mass index (BMI), and study site. Significant proteins were identified using a Bonferroni-corrected p-value <0·05 based on a t-test for the regression coefficient. All analyses were stratified by smoking status (ever-vs. never-smokers). In a sensitivity analysis, we additionally adjusted for chronic kidney disease and lung disease.

## RESULTS

### MGB cohort characteristics

Our primary MGB study cohort consisted of 12,550 patients who had ever smoked cigarettes (mean age 62·4 [SD 6·8], 48·9% male, 90·6% White), and 15,298 patients who had never smoked (mean age 63·0 [SD 8·1], 42·8% male, 85·8% White) (Table 1). In this cohort, the 6-year COPD incidence was 12·4% (1562/12550) in ever-smokers and 3·8% (580/15298) in never-smokers. Cohort characteristics were largely similar to the cohort in which the CXR-Lung-Risk model was developed (Supplementary Table 2); however, the smokers in the current cohort had a milder smoking history (mean 15·9 pack-years vs 35·5).

**Table 1.** Cohort characteristics for Mass General Brigham patients.

|  | **Ever-smoker group** | **Non-smoker group** |
| --- | --- | --- |
| N | 12550 | 15298 |
| Age, mean (SD) | 62.4 (6.8) | 63.0 (8.1) |
| Male sex (%) | 6135 (48.9) | 6550 (42.8) |
| Race (%) |  |  |
| Asian | 189/11569 (1.6) | 707/13686 (5.2) |
| Black | 874/11569 (7.6) | 1174/13686 (8.6) |
| Other Race | 19/11569 (0.2) | 57/13686 (0.4) |
| White | 10487/11569 (90.6) | 11748/13686 (85.8) |
| Hispanic ethnicity (%) | 349/12338 (2.8) | 547/13356 (4.0) |
| Recent dyspnea (%) | 4519 (36.0) | 6503 (42.5) |
| Salbutamol use (%) | 3723 (29.7) | 3385 (22.1) |
| Antibiotic use (%) | 2378 (18.9) | 2180 (14.3) |
| Current smoker (%) | 2366 (18.9) | - |
| Smoking pack-years, mean (SD) | 15.9 (20) | - |
| CXR-Lung-Risk, mean (SD) | 50.9 (5.7) | 49.3 (5.9) |
| TargetCOPD score mean (SD) | 0.14 (0.1) | 0.06 (0.05) |
| CXR report findings (%) |  |  |
| Lung opacity | 2631 (21.0) | 3024 (19.8) |
| Atelectasis | 1374 (10.9) | 1565 (10.2) |
| Pneumothorax | 279 (2.2) | 250 (1.6) |
| Pneumonia | 1194 (9.5) | 1397 (9.1) |
| Edema | 484 (3.9) | 481 (3.1) |
| Consolidation | 310 (2.5) | 380 (2.5) |
| Lung lesion | 1357 (10.8) | 1830 (12.0) |
| Prevalent asthma (%) | 1338 (10.7) | 2465 (16.1) |
| 6-year COPD rate (%) | 1562 (12.4) | 580 (3.8) |
| 6-year all-cause mortality (%) | 944/12219 (7.7) | 1978 (13.0) |

COPD, chronic obstructive pulmonary disorder.

### CXR-Lung-Risk model discrimination

We first evaluated the discrimination of CXR-Lung-Risk and baseline approaches to predict 6-, 3-, and 1-year COPD incidence. CXR-Lung-Risk significantly improved the AUC for 6-year COPD incidence beyond the TargetCOPD clinical risk calculator both among ever-smokers (CXR-Lung-Risk + TargetCOPD AUC: 0·73 [95% CI: 0·72-0·74] vs. TargetCOPD AUC: 0·66 [95% CI: 0·65-0·68], p<0·01) and never-smokers (CXR-Lung-Risk + TargetCOPD AUC: 0·70 [95% CI: 0·67-0·72] vs. TargetCOPD AUC: 0·60 [95% CI: 0·57-0·62], p<0·01) (Table 3). Similar results were seen for 3- and 1-year outcomes.

**Table 2.**
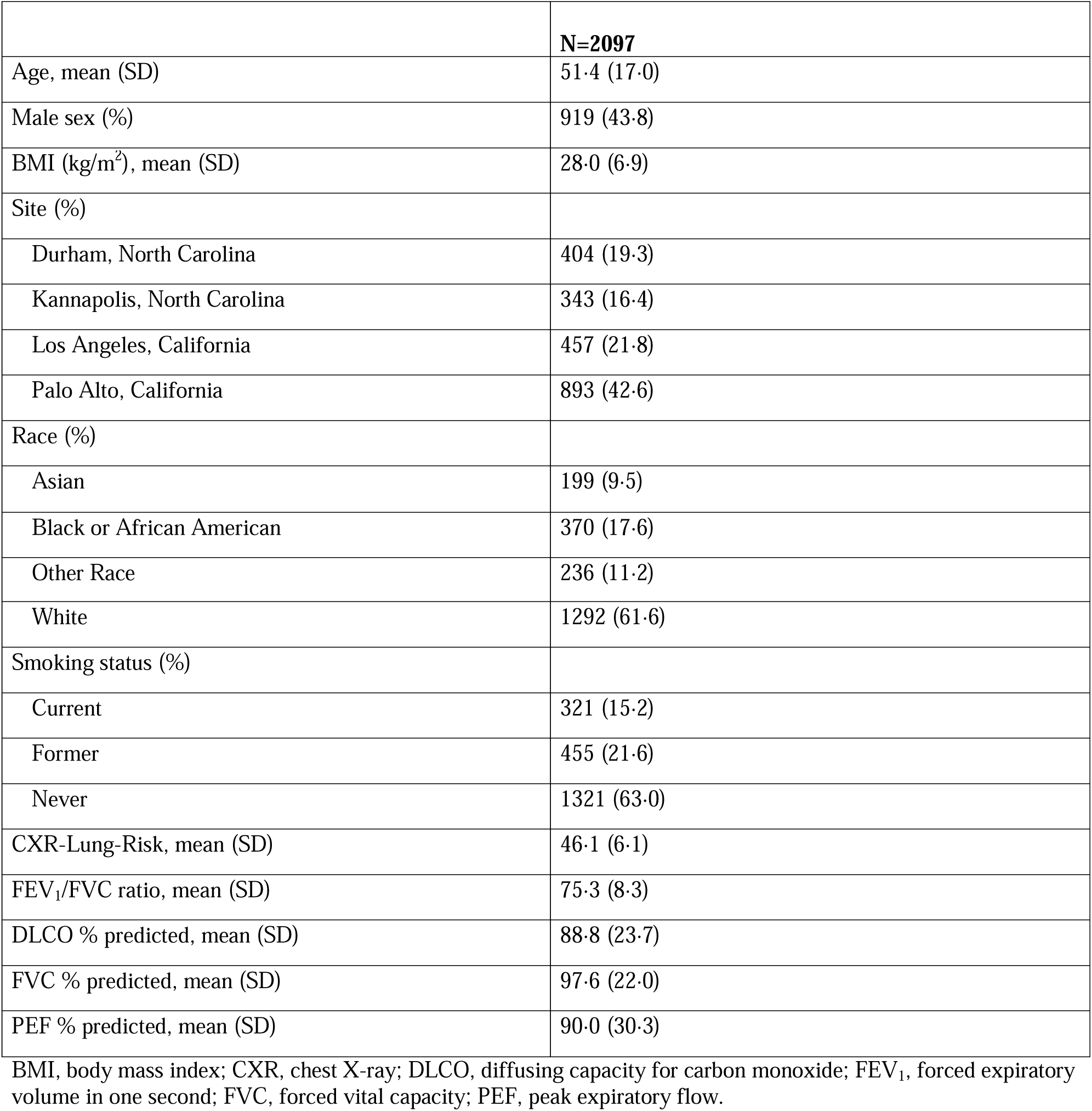
Cohort characteristics for Project Baseline Health Study participants.

**Table 3.** Discrimination for 6-year, 3-year, and 1-year incident chronic obstructive pulmonary disease (COPD) by baseline, CXR-Lung-Risk, and TargetCOPD models in ever-smokers and never-smokers.

| **Model** | **Ever-smoker group**<br>**AUC [95% CI]** | **Non-smoker group**<br>**AUC [95% CI]** |
| --- | --- | --- |
| **6-year incident COPD** |  |  |
| Age, Sex | 0.53 [0.52–0.55] | 0.60 [0.57–0.62] |
| Age, Sex, Smoking Status | 0.64 [0.63–0.66] | N/a |
| TargetCOPD | 0.66 [0.65–0.68] | 0.60 [0.57–0.62] |
| CXR-Lung-Risk | 0.67 [0.66–0.69] | 0.66 [0.64–0.68] |
| CXR-Lung-Risk + TargetCOPD | 0.73 [0.72–0.74] | 0.70 [0.67–0.72] |
| **3-year incident COPD** |  |  |
| Age, Sex | 0.54 [0.52–0.56] | 0.59 [0.56–0.62] |
| Age, Sex, Smoking Status | 0.65 [0.63–0.67] | N/a |
| TargetCOPD | 0.68 [0.66–0.70] | 0.58 [0.55–0.61] |
| CXR-Lung-Risk | 0.67 [0.66–0.69] | 0.65 [0.63–0.68] |
| CXR-Lung-Risk + TargetCOPD | 0.74 [0.73–0.76] | 0.69 [0.66–0.71] |
| **1-year incident COPD** |  |  |
| Age, Sex | 0.53 [0.50–0.56] | 0.60 [0.57–0.64] |
| Age, Sex, Smoking Status | 0.63 [0.61–0.66] | N/a |
| TargetCOPD | 0.68 [0.65–0.70] | 0.56 [0.52–0.60] |
| CXR-Lung-Risk | 0.67 [0.66–0.69] | 0.67 [0.64–0.70] |
| CXR-Lung-Risk + TargetCOPD | 0.73 [0.72–0.74] | 0.69 [0.66–0.73] |

\*P<0.05.
AUC, area under the receiver operating characteristic curve; CI, confidence interval.

### Ordinal CXR-Lung-Risk model categories and incident COPD

We tested the association of ordinal CXR-Lung-Risk groups with incident COPD (Supplementary Table 3). In ever-smokers, a graded association with incident COPD was observed across moderate (aHR: 1·7 [95% CI: 1·5-1·9]) and high (aHR: 3·4 [95% CI: 2·9-3·9]) CXR-Lung-Risk groups compared to the low-risk group, after adjusting for risk factors and radiologist findings on the CXR report (Fig. 2a).

**Figure 2.**
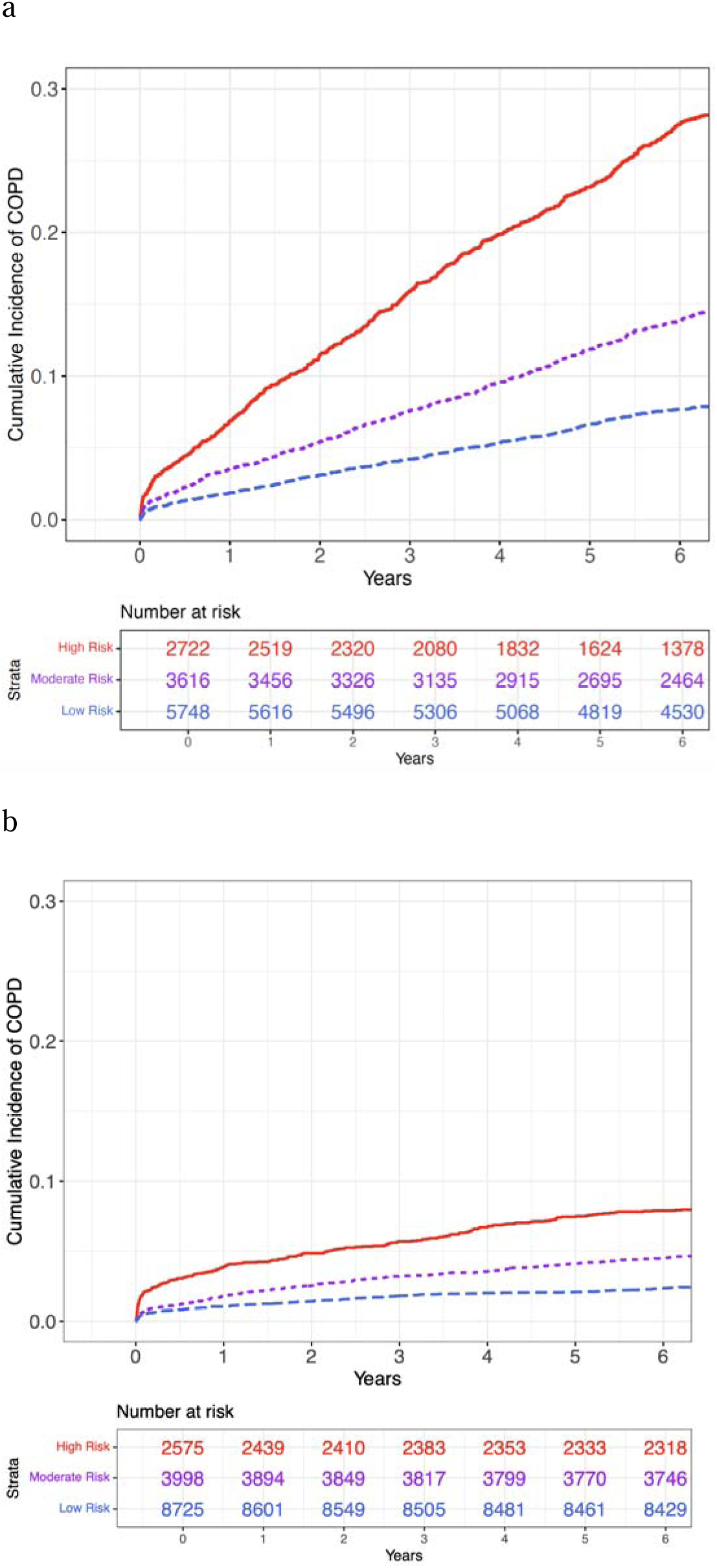
Cumulative incidence of chronic obstructive pulmonary disease (COPD) by CXR-Lung-Risk ordinal categories in (a) ever-smokers and (b) never-smokers.

A similar pattern was observed in never-smokers, with a higher risk of incident COPD in the moderate (aHR: 1·6 [95% CI: 1·3-1·9]) and high (aHR: 2·4 [95% CI: 1·9-3·0]) CXR-Lung-Risk groups compared to the low-risk group after adjustment (Fig. 2b).

### COPD rates at binary thresholds

We compared the rates of incident COPD among patients at high risk according to CXR-Lung-Risk and those at high risk according to the TargetCOPD score using the published 7·5% risk threshold (Table 4).^19^ We found that the two risk models were complimentary, with patients at high risk by both models having a 28·4% and 11·5% rate of 6-year COPD in the ever-smoker group and never-smoker group, respectively. Patients at low risk by both approaches had a 4·7% and 2·2% rate of 6-year COPD in the ever-smoker group and never-smoker group, respectively. Similar results were found for 3- and 1-year incident COPD (Supplementary Table 4).

**Table 4.** Rates of 6-year chronic obstructive pulmonary disease (COPD) in ever- and never-smokers by CXR-Lung-Risk and TargetCOPD binary high-risk groups.

|  | **TargetCOPD**<br>**>7.5%** | **TargetCOPD**<br>**≤7.5%** | **Total** |
| --- | --- | --- | --- |
| **Ever-smokers** |  |  |  |
| CXR-Lung-Risk High | 549/1936 (28.4%) | 128/934 (13.7%) | 677/2870 (23.6%) |
| CXR-Lung-Risk Not High | 703/5832 (12.0%) | 182/3848 (4.7%) | 885/9680 (9.1%) |
| **Total** | 1252/7768 (16.1%) | 310/4782 (6.5%) | 1562/12550 (12.4%) |
| **Never-smokers** |  |  |  |
| CXR-Lung-Risk High | 64/558 (11.5%) | 136/2017 (6.7%) | 200 / 2575 (7.8%) |
| CXR-Lung-Risk Not High | 165/2990 (5.5%) | 215/9733 (2.2%) | 380 / 12723 (3.0%) |
| **Total** | 229/3548 (6.4%) | 351/11750 (3.0%) | 580/15298 (3.8%) |

### Association of CXR-Lung-Risk with pulmonary function tests and proteomics in the Project Baseline Health Study

For additional insight into the biological basis of the association between CXR-Lung-Risk and incident COPD, we tested whether CXR-Lung-Risk was associated with quantitative measures of pulmonary function and plasma protein abundance using data from 2,097 individuals with a CXR image as part of the PBHS study (mean age 62·3± 6·8, 48·6% male, 82·7% non-Hispanic White; Table 2).

CXR-Lung-Risk was negatively correlated with all pulmonary function measures tested (FEV_1_/FVC, diffusing capacity for carbon monoxide [DLCO], FEV_1_, PEF) in ever-smokers (R^2^: 6%-22%) and in never-smokers, although to a lesser extent (R^2^: 1%-8%) (Supplementary Fig 1). These relationships persisted after adjusting for further covariates in linear regression, including age, sex, BMI, and study site. With every 1 standard deviation (SD) increase in CXR-Lung-Risk (∼6 years), a 2·4%–5·3% decrease in pulmonary function test performance was observed for ever-smokers (p<0·05 for all comparisons) after adjustment. Associations were attenuated in never-smokers, although the FEV_1_/FVC ratio (0·8% [0·3%-1·3%] per SD of CXR-Lung-Risk) and DLCO % predicted (1·8% [0·1%-3·6%]) were associated after adjustment (both p<0·05). When adjusting for the presence of any type of lung disease, the effect sizes were attenuated but remained significant except for PEF (Supplementary Fig. 2).

In proteomic analyses across 289 plasma proteins, we found that two proteins had a significant positive relationship with the CXR-Lung-Risk score (Bonferroni-adjusted p-value <0·05): SCGB3A2 (secretoglobin family 3A member 2) and LYZ (lysozyme) (Fig. 3). This was robust to adjustment for history of lung disease or chronic kidney disease (Supplementary Fig. 3a). In a stratified analysis by smoking status, SFTPB (surfactant protein B) and LRG1 (leucine-rich α-2 glycoprotein 1) were significantly associated with CXR-Lung-Risk in ever-smokers, whereas LYZ and SCGB3A2 had similar effect sizes but did not have a significant relationship with

**Figure 3.**
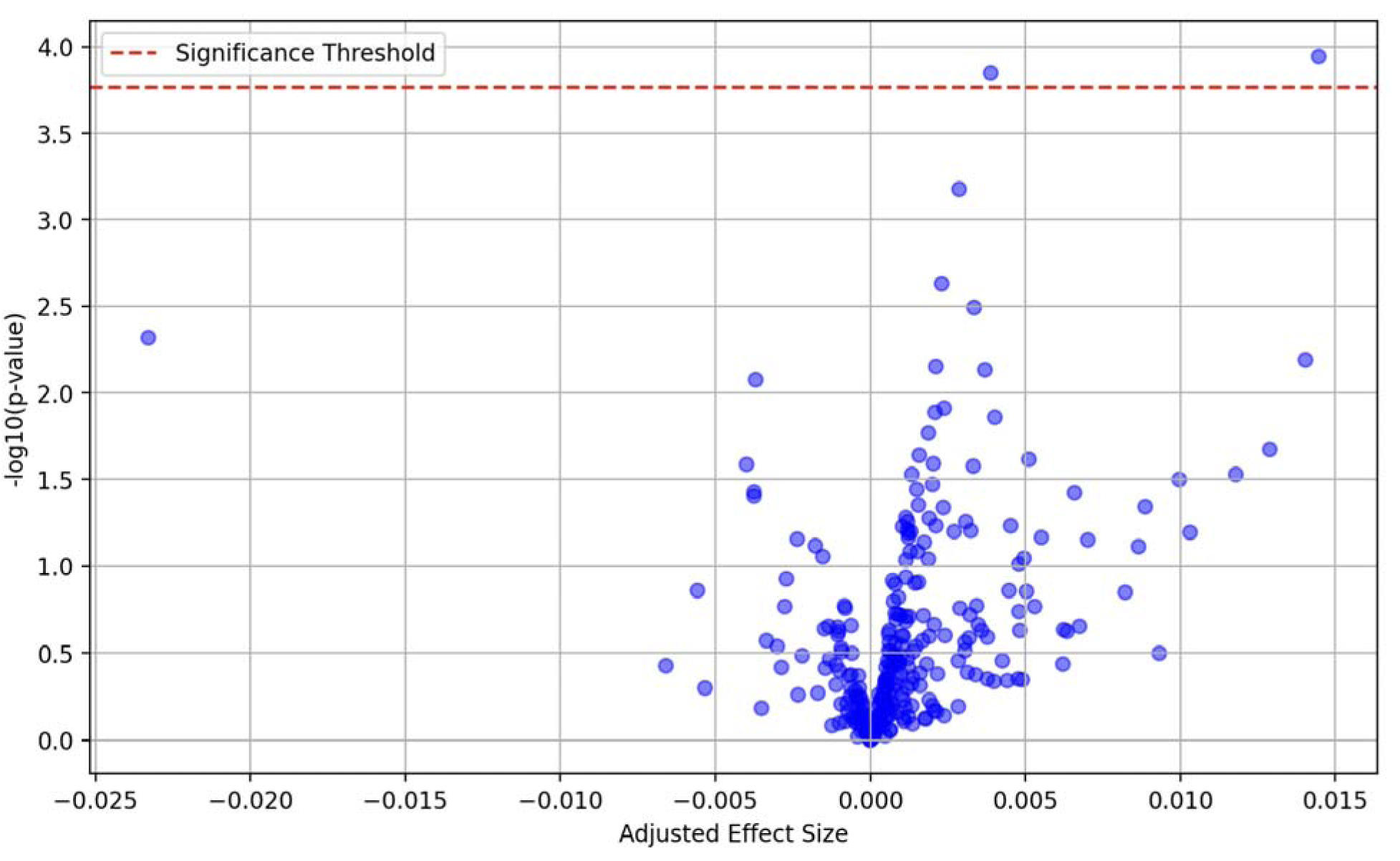
Association between CXR-Lung Risk model and plasma protein abundance in the Project Baseline Health Study (PBHS). All estimates are adjusted for age, sex, body mass index (BMI), study site, smoking status, and frequency of smoking if the patient has smoked (every day vs some days).

CXR-Lung-Risk (Supplementary Fig. 3b). For never-smokers, LYZ had a similar adjusted effect size as in the full analysis with all patients; however, the association of SCGB3A2 and SFTPB with CXR-Lung-Risk was attenuated in never-smokers (Supplementary Fig. 3c).

## DISCUSSION

Chest radiographs are a basic, first-line test for respiratory symptoms; however, these images are non-diagnostic for COPD. We hypothesized that an AI model, CXR-Lung-Risk, could extract “hidden” information from the CXR image to identify individuals at high risk for incident COPD. Our major findings were 1) CXR-Lung-Risk predicted 6-year incident COPD with complimentary value to a clinical risk score in ever-smokers (ΔAUC=0·07 for combined vs. clinical score alone) and never-smokers (ΔAUC=0·10), 2) Patients at the highest risk of incident COPD according to the AI model had high rates of 6-year COPD (ever-smokers: 23·6%; never-smokers: 7·8%) and 3) Higher CXR-Lung-Risk was associated with poor performance on PFTs and with plasma protein concentrations with known relationships to lung function.

Despite significant advances in establishing diagnostic criteria for COPD,^27^ it is estimated that half of COPD cases remain undiagnosed. Potential reasons include the lack of spirometry use, lack of public awareness of symptoms and risk factors, misinterpretation of spirometry results in younger adults and the elderly, and manifestation of similar respiratory symptoms in patients with comorbidities.^9,10^ Since persistent respiratory symptoms are required to diagnose COPD, routine screening of asymptomatic adults is not a recommended, nor feasible solution. Instead, targeted case finding via patient questionnaires or risk factor-based scores may be a more effective solution to identify individuals at high risk who are likely to be diagnosed with COPD upon spirometric testing.^28^

The CXR-Lung-Risk model presented here may improve early identification of individuals at high risk for COPD through opportunistic screening of existing routine CXRs in the EMR. The CXR-Lung-Risk model could scan the electronic medical record to identify patients with CXRs already administered during routine care stored in electronic Picture Archiving and Communication Systems (PACS) systems and estimate COPD risk. For patients with a high estimated risk, the system could alert their care team that they may be at high risk for COPD. In our current study, ever-smokers in the highest risk group according to CXR-Lung-Risk had a 23·6% rate of incident COPD over 6 years (7·8% in never-smokers). Additionally, the CXR-Lung-Risk model may be recognizing signs consistent with undiagnosed COPD, as 6·5% and 3·8% of ever- and never-smokers at high predicted risk had COPD diagnosed within 1 year of the CXR, respectively, suggesting that 16–25 individuals need to be screened to detect one undiagnosed case of COPD. Potential next steps for patients at high estimated risk include: 1) sending the patient a respiratory symptom questionnaire, 2) conducting surveillance for signs and symptoms of incident COPD, or 3) performing diagnostic spirometry.

Adding the CXR-Lung-Risk model to the TargetCOPD clinical risk score increased the discrimination for 6-year incident COPD in ever-smokers (ΔAUC=0·07) and never-smokers (ΔAUC=0·10). This suggests that further performance improvements are possible when combining the CXR image with prevalent risk factors and smoking history from the medical record, which will be explored in future work.

A common concern with AI-based approaches is their lack of interpretability or “black-box” nature.^29^ In our previous study, we showed that the CXR-Lung-Risk outputs were associated with prevalent risk factors including age, sex, obesity, smoking status, smoking pack-years, history of cardiovascular disease, and the presence of emphysema, fibrosis, and lung opacities on the CXR image. In this study, we found strong associations between the CXR-Lung-Risk output with lower lung function and abundance of three lung-related proteins in a community cohort from the PBHS study. DLCO values and the FEV_1_/FVC ratio were negatively associated with CXR-Lung-Risk with a stronger effect observed in smokers. The former is a marker of nearly all lung diseases and the latter is a marker of obstructive lung diseases such as COPD and asthma.^30^ All PFTs (PEF, FEV_1_/FVC ratio, and DLCO) were negatively associated with CXR-Lung-Risk in smokers.

To gain further insight into potential biological mechanisms of high CXR-Lung-Risk, we tested associations between our risk score and plasma protein concentrations. We found positive associations of SCGB3A2 and LYZ concentrations with CXR-Lung-Risk and a strong association between SFTPB with CXR-Lung-Risk, although non-significant. SCGB3A2 is a cytokine molecule only expressed in the lungs by the bronchiolar club cells.^31^ It has been shown to be protective against various lung disease processes including inflammation, fibrosis, and malignancy.^31^ Additionally, SCGB3A2 has been shown to be associated with asthma and COPD.^32,33^ SFTPB is a lung surfactant protein only expressed in the lungs. Higher plasma expression levels of SFTPB have been observed in patients with lung disease and decreased lung function potentially due to increased lung permeability that enables the transit of surfactant into the blood.^34,35^ LYZ is an enzyme that primarily degrades bacterial cell walls. It is expressed in the lungs, plasma, stomach, and salivary glands.^36^ LYZ was found to have higher expression levels in the lungs in the setting of COPD and idiopathic pulmonary fibrosis.^37,38^ Additionally, LYZ has been specifically linked to the development of pulmonary emphysema.^36^ These analyses suggest that CXR-Lung-Risk may be picking up imaging signs specific to biological pathways of reduced lung function.

Limitations of this study should be considered. The primary analysis was conducted in patients having routine chest radiography at a single hospital system in Boston, Massachusetts, and most were non-Hispanic White individuals. Future studies need to test this model in more diverse populations and other geographic locations, especially as COPD may have a heterogenous etiology across racial/ethnic groups.^39^ Although the CXR-Lung-Risk model accurately identified individuals at high risk for incident COPD, it is unclear whether this will improve early detection; this needs to be tested in prospective trials.^40^ We chose to use radiographs taken in 2013–2014 to ensure a 6-year follow-up period, but trends in COPD diagnosis and prevalence may have changed. A criticism of risk prediction approaches is that they tend to identify older and frailer individuals as high-risk; this problem of overdiagnosis needs to be addressed in a prospective trial.^41^

In this study, we externally validated the CXR-Lung-Risk model, an open-source AI tool, for the identification of patients at high risk of COPD based on a routine chest radiograph image from the EMR. CXR-Lung-Risk predictions were associated with pulmonary function testing and with plasma proteins indicative of lung health. Future research will test whether implementation of this model can improve the high undiagnosed case rate of COPD across diverse populations.

## CONTRIBUTORS

Study design: S.D.J, V.R., M.T.L .; code design, implementation and execution: SDJ, JC, ASW, AS, VKR; acquisition, analysis or interpretation of data: SDJ, JC, VKR, MTL, JW, HJWLA, DC; writing of the manuscript: SDJ, JC, VKR; critical revision of the manuscript for important intellectual content: all authors; statistical analysis: SDJ, JC, VKR; study supervision: VKR, MTL. All authors had full access to all the data in the study and had final responsibility for the decision to submit for publication. VKR and SDJ have accessed and verified the data.

## DATA SHARING STATEMENT

The deidentified PBHS data corresponding to this study are available upon request for the purpose of examining its reproducibility. Requests are subject to approval by PBHS governance. Due to institutional policy to protect patient privacy, MGB data cannot be shared.

## ACKNOWLEDGMENTS

The Baseline Health Study and this analysis were funded by Verily Life Sciences, San Francisco, California. The authors wish to thank Project Baseline Health Study participants and study sites. The authors would also like to thank Brooke Walker, MS, Duke Clinical Research Institute, who provided editorial support. Ms. Walker did not receive compensation for her contributions, apart from her employment at the institution in which this study was conducted.

