## Supplementary Material for "Leveraging Deep Learning of Chest Radiograph Images to Identify Individuals at High Risk for Chronic Obstructive Pulmonary Disease"

| Supplementary Methods | 2 |
| --- | --- |
| Supplementary Table 1. ICD-9 and -10 codes used to identify chronic obstructive pulmonary disease cases^30,49^ | 4 |
| Supplementary Table 2. Comparison of current validation cohort with CXR-Lung-Risk model development cohort | 5 |
| Supplementary Table 3. Association of ordinal CXR-Lung-Risk groups with incident chronic obstructive pulmonary disease in ever- (left) and never-smokers (right) | 6 |
| Supplementary Table 4. Rates of 3-year (left)and 1-year (right) incident chronic obstructive pulmonary disease (COPD) in ever- (top) and never-smokers (bottom) by CXR-Lung-Risk and TargetCOPD binary high-risk groups | 7 |
| Supplementary Figure 1. CONSORT diagram for never- (left) and ever- (right) smokers in the Massachusetts General Brigham (MGB) cohort | 8 |
| Supplementary Figure 2. Association between pulmonary function tests and CXR-Lung-Risk model in ever-smokers (blue) and never-smokers (orange) in the Project Baseline Health Study (PBHS). All estimates are adjusted for age, sex, body mass index (BMI), study site, and frequency of smoking if the patient has smoked (every day vs some days). | 9 |
| Supplementary Figure 3. Association between pulmonary function tests and CXR-Lung-Risk in ever- (blue) and never- (orange) smokers in the Project Baseline Health Study controlled for lung disease | 10 |
| Supplementary Figure 4. Association of CXR-Lung-Risk with plasma protein concentrations (a) after adjustment for lung and kidney disease, (b) in ever-smokers, and (c) in never-smokers | 12 |

**Supplementary Methods**

**CXR-Lung-Risk model**

The model was an ensemble of 20 convolutional neural networks (CNNs) with varied architectures (inceptionv4, resnet34, tiny) and hyperparameters. The output of these models was combined using LASSO regression, with 13 out of 20 models included in the final ensemble. Radiographs from the Prostate, Lung, Colorectal, and Ovarian (PLCO) Cancer Screening Trial were provided as scanned films in Tagged Image File Format (.tif) with protected health information redacted using black pixels. PLCO radiographs were converted to Portable Network Graphics (.png) format using ImageMagick, v6.8.9-9. During the training phase, images were resized to 224 pixels on the short-axis and randomly cropped to 224×224 pixels before use as an input in the model. This random cropping served as a method of data augmentation. Additionally, augmentations applied during training involved mixup data augmentation, allowing up to 20 degrees of random rotation, up to 20% zoom in/out, and up to 40% adjustments in brightness/contrast. The model was trained using the ADAM optimizer and a mean-squared error loss function. The number of training epochs, ranging from 40-70, was randomly chosen for each of the 20 models. Training was conducted on an Ubuntu Linux workstation with an AMD 3960×24-core CPU (128 GB RAM) and a single NVIDIA RTX A6000 GPU (48 GB GPU RAM). The development was carried out using fastai, v2.5.3; PyTorch, v1.10; and CUDA, v11.2. The full source code can be accessed through this link: https://aim.hms.harvard.edu/cxr-lungrisk.

**Convolutional neural network to predict lung-related mortality from chest X-ray (CXR-Lung-Risk)**

We previously developed a convolutional neural network (CXR-Lung-Risk) to predict lung-related mortality from chest X-rays (CXRs).^25^ In brief, this model was developed by estimating chronological age using public CXR datasets for ‘pre-training’, then using this Stage I model as a starting point for estimating biological age using the PLCO training data. We defined the biological age labels based on an individuals’ expected risk of long-term lung-related mortality using the formula below:

*BA = CA + (E-D)*

where *BA* is the biological age label; *CA* is the chronological age at the time of chest x-ray; *E* is the expected age-at-death according to the United States (U.S.) Social Security Administration; and *D* is the actual, observed age at death for those who died of lung disease and estimated age-at lung-related death based on a survival model for those who did not die of lung disease during follow-up. In the original study, this model was validated in two held-out testing datasets from the PLCO trial and the National Lung Screening Trial (NLST) not used during model development.

**Chest radiograph preprocessing steps**

Images were acquired in Digital Imaging Communications in Medicine (DICOM) format and transformed into .png format using identical image pre-processing procedures applied in the creation of a previously developed pipeline by our group, called “CXR-LC” (<https://github.com/circ-ml/CXR-LC>).^24^ A deep learning model, designed to predict the view position from the radiograph image, identified low-quality CXR images (<https://github.com/circ-ml/CXR-View>). Images were discarded if the model had less than 80% confidence that the image was a posterior-anterior CXR.

The content of the radiologists' reports for the CXRs was retrieved from the Electronic Medical Record (EMR). Detection of lung nodules and other radiographic observations was performed using the CheXpert-labeler software, which is an open-source tool designed to identify radiographic findings from free-text reports.^48^

### **Proteomics data**

### *Data generation*

In brief, plasma proteins were prepared through Verily Life Sciences’s proteomics pipeline, utilizing robotic liquid handling and validated plasma preparation kits to achieve high-throughput processing, consistency, and scale. For each plasma sample, 2 microliters were denatured with trypsin/Lys-C protease and the subsequent peptides were desalted and dried down in a vacuum concentrator. Dried pellets were dissolved in 40 microliters of 0·1% (v/v) formic acid, then peptide concentrations were normalized to 1 microgram per microliter and combined with iRT standard peptides (1:20 v/v). Next, 5 micrograms of each sample was randomly injected in duplicate onto a customized microflow high-resolution liquid chromatography-mass spectrometry (LC-MS) setup. Mass spectra were acquired in data-independent acquisition (DIA) mode for accurate and reproducible quantification. Raw data files were saved locally and on the cloud for downstream analysis.

#### *Peptide abundance quantification*

According to the experimental design, each sample is run as two technical replicates for each batch. If the instrument performance is degrading during a batch, more than two replicates are processed. Custom code is used, unless specified otherwise.

Mass spectra are stored as proprietary ThermoFisher .raw files. The spectra are analyzed to infer peptide abundances through several processing steps:

1. The .raw files are converted to .mzML format using msconvert from ProteoWizard (<https://proteowizard.sourceforge.io/>). In addition, .mzML files are centroid-normalized for compatibility.
2. Dia-NN, v1.8.1 (<https://github.com/vdemichev/DiaNN>), is executed in library-free mode to generate preliminary peptide abundances. The plasma proteome from the Human PeptideAtlas ([http://www.peptideatlas.org/builds/human/plasma](http://www.peptideatlas.org/builds/human/plasma/)) is used to restrict the search space to plasma protein sequences. Dia-NN is executed with the following options:

–f file.mzML –fasta PeptideAtlas.fasta –fasta-search –gen-spec-lib –gen-fr-restriction –mass-acc-ms1 5 –mass-acc 15 –min-pr-mz 400 –max-pr-mz 880 –window 6 –restrict-fr –no-fr-selection –no-norm –no-maxlfq –individual-reports

3. A subset of samples is analyzed jointly to generate the spectral library with Dia-NN. Specifically, the first available replicate of each sample is selected from each participant’s entry visit. Dia-NN parameters are the same as in the previous step.
4. Each replicate sample is reprocessed independently using the newly generated spectral library. Dia-NN parameters are the same as in the previous steps, except now the parameters related to the library-free search, namely –fasta PeptideAtlas.fasta –fasta-search –gen-spec-lib –gen-fr-restriction, are replaced with –lib library.tsv –no-prot-inf.

#### *Protein abundance quantification*

Peptide abundances quantified with Dia-NN are filtered, normalized, and aggregated to protein abundances through several processing steps:

1. Quality metrics are computed and used to filter samples and precursors used for the analyses. Specifically:
   a) When a batch is repeated, only the latest batch is used for analysis.
   b) Only replicates with more than 2500 precursors are used for analysis.
   c) Only samples with at least two valid technical replicates are used for analysis. In the case of more than two technical replicates per sample, the first two are used for analysis.
   d) Proteins need to have a Dia-NN library q-value <0·01.
   e) Proteins need to have a Dia-NN q-value <0.05 in both replicates in at least 100 samples.
   f) Precursors need to be reproducible between replicates, with a coefficient of variation <0·2 for all samples.
   g) Microbial proteins, contaminants, and Ig variable chain proteins are not included in downstream analyses.
2. Mass spectrometer performance may degrade as samples are loaded during a batch run. This may result in precursor expression drift as a function of when the sample was loaded on the instrument. A polynomial regression for the log-transformed precursor expression is fit on the run order for each precursor in each batch, and predictions are regressed out to the median to adjust for temporal bias.
3. Non-log-transformed normalized precursor quantities are summed up to compute protein abundances within each replicate.
4. Protein quantities are log-transformed again and averaged between technical replicates to obtain protein quantities at the sample level. Missing values in one replicate are imputed with values from the other replicate.

Finally, averaged protein quantities are corrected for batch effects using a python implementation of the ComBat method (<https://github.com/epigenelabs/pyComBat>).

**Supplementary Table 1. ICD-9 and -10 codes used to identify chronic obstructive pulmonary disease cases^30,49^**

| **Codes** | **ICD** |
| --- | --- |
| 491 | 9 |
| 492 | 9 |
| 496 | 9 |
| J41 | 10 |
| J42 | 10 |
| J43 | 10 |
| J44 | 10 |

ICD, International Classification of Diseases.

**Supplementary Table 2. Comparison of current validation cohort with CXR-Lung-Risk model development cohort**

|  | **Ever-smoker group (MGB)** | **Non-smoker group (MGB)** | **PLCO ever-smokers (development cohort)** | **PLCO non-smokers (development cohort)** |
| --- | --- | --- | --- | --- |
| N | 12550 | 15298 | 23365 | 18666 |
| Age, mean (SD) | 62·4 (6·8) | 63·0 (8·1) | 62·2 (5·3) | 62·6 (5·5) |
| Male sex (%) | 6135/12550 (48·9%) | 6550/15298 (42·8%) | 14075/23365 (60·2%) | 7671/18666 (41·1%) |
| Race (%) |  |  |  |  |
| Asian | 189/11569 (1·6%) | 707/13686 (5·2%) | 863/22882 (3·8%) | 1039/18349 (5·7%) |
| Black | 874/11569 (7·6%) | 1174/13686 (8·6%) | 1488/22882 (6·5%) | 990/18349 (5·4%) |
| Other Race | 19/11569 (0·2%) | 57/13686 (0·4%) | 192/22882 (0·8%) | 155/18349 (0·8%) |
| White | 10487/11569 (90·6%) | 11748/13686 (85·8%) | 20339/22882 (88·9%) | 16165/18349 (88·1%) |
| Hispanic ethnicity (%) | 349/12338 (2·8%) | 547/13356 (4·0%) | 508/22766 (2·2%) | 334/17952 (1·9%) |
| Smoking pack-years, mean (SD) | 15·9 (20·0) | NA | 35.2 (29.0) | NA |

MGB, Massachusetts General Brigham; PLCO, Prostate, Lung, Colorectal, and Ovarian Cancer Screening Trial.

**Supplementary Table 3. Association of ordinal CXR-Lung-Risk groups with incident chronic obstructive pulmonary disease in ever- and never-smokers**

|  | **Ever-smokers**  **(N= 12550)** | | | | **Never-smokers**  **(N= 15298)** | | | |
| --- | --- | --- | --- | --- | --- | --- | --- | --- |
| **CXR-Lung-Risk** | **6-year COPD rate (%)** | **Per 1000 person-years (95% CI)** | **Adj HR* (95% CI)** | **P-value** | **6-year COPD rate (%)** | **Per 1000 person-years (95% CI)** | **Adj HR* (95% CI)** | **P-value** |
| Low | 421/5923 (7·1%) | 12·4 (11·3–13·6) | Ref |  | 202/8725 (2·3%) | 4·0 (3·4–4·5) | Ref |  |
| Moderate | 464/3757 (12·4%) | 22·7 (20·6–24·7) | 1·7 (1·5–1·9) | <0·001 | 178/3998 (4·5%) | 7·7 (6·6–8·9) | 1·6 (1·3–1·9) | <0·001 |
| High | 677/2870 (22·0%) | 48·0 (44·4–51·6) | 3·4 (2·9–3·9) | <0·001 | 200/2575 (7·8%) | 14·0 (12·0–15·9) | 2·4 (1·9–3·0) | <0·001 |
| Total | 1562/12550 (12·5%) | 22·8 (21·7–24·0) |  |  | 580/15298 (3·8%) | 6·6 (6·0–7·1) |  |  |

*Hazard ratio (HR) adjusted for age, sex, smoking status, recent dyspnea, number of salbutamol prescriptions, whether the patient is on salbutamol, prescription of antibiotics, history of asthma, and lung opacity, atelectasis, pneumothorax, pneumonia, edema, consolidation, or lung lesion noted on the chest X-ray (CXR) report.

**Supplementary Table 4. Rates of 3-year (left) and 1-year (right) incident chronic obstructive pulmonary disease (COPD) in ever- (top) and never- (bottom) smokers by CXR-Lung-Risk (rows) and TargetCOPD (columns) binary high-risk groups**

|  | **3-year incident COPD rates** | | **1-year incident COPD rates** | |
| --- | --- | --- | --- | --- |
|  | **TargetCOPD**  **>7·5%** | **TargetCOPD**  **<=7·5%** | **TargetCOPD**  **>7·5%** | **TargetCOPD**  **<=7·5%** |
| **Ever-smokers** |  |  |  |  |
| CXR-Lung-Risk High | 347/1936 (17·9%) | 75/934 (8·0%) | 152/1936 (7·8%) | 34/934 (3·6%) |
| CXR-Lung-Risk Not High | 421/5832 (7·2%) | 86/3848 (2·2%) | 206/5832 (3·5%) | 27/3848 (0·7%) |
| **Never-smokers** |  |  |  |  |
| CXR-Lung-Risk High | 40/558 (7·2%) | 105/2017 (5·2%) | 22/558 (3·9%) | 77/2017 (3·8%) |
| CXR-Lung-Risk Not High | 125/2990 (4·2%) | 159/9733 (1·6%) | 69/2990 (2·3%) | 93/9733 (0·9%) |

**Supplementary Figure 1: CONSORT diagram for never- (left) and ever- (right) smokers in the Massachusetts General Brigham (MGB) cohort**

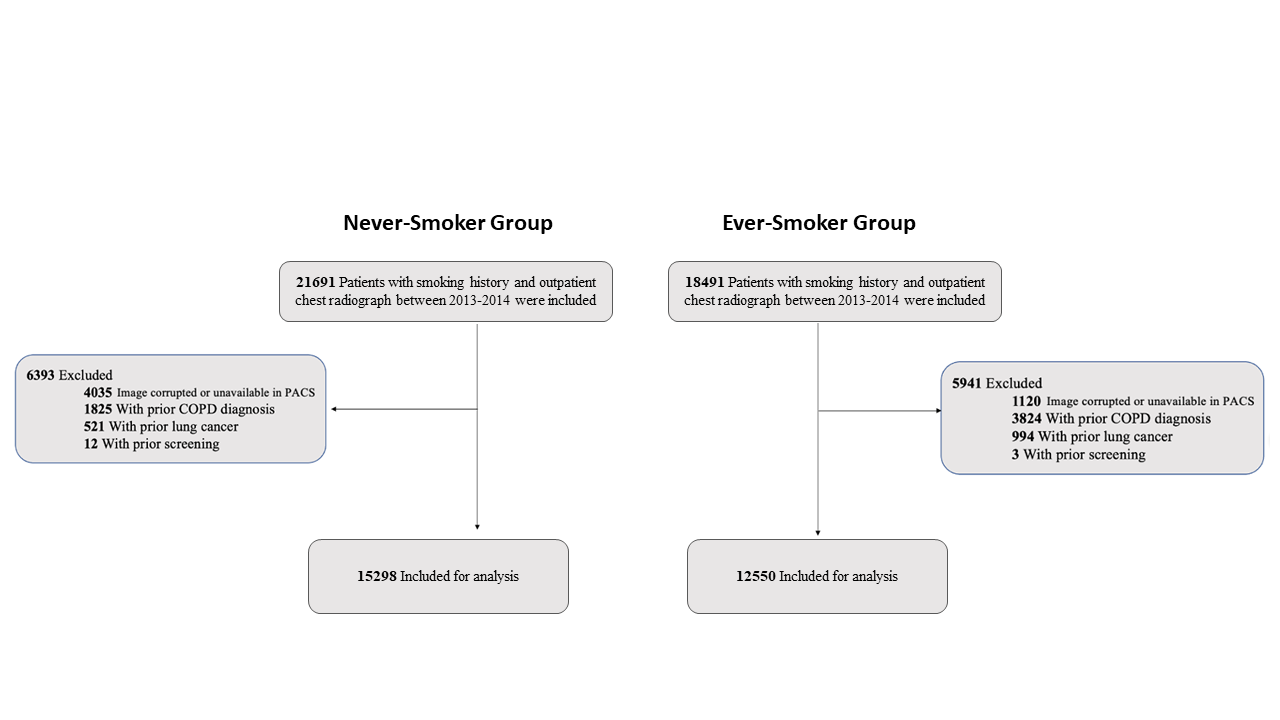

**Supplementary Figure 2. Association between pulmonary function tests and CXR-Lung-Risk model in ever-smokers (blue) and never-smokers (orange) in the Project Baseline Health Study (PBHS). All estimates are adjusted for age, sex, body mass index (BMI), study site, and frequency of smoking if the patient has smoked (every day vs some days).**

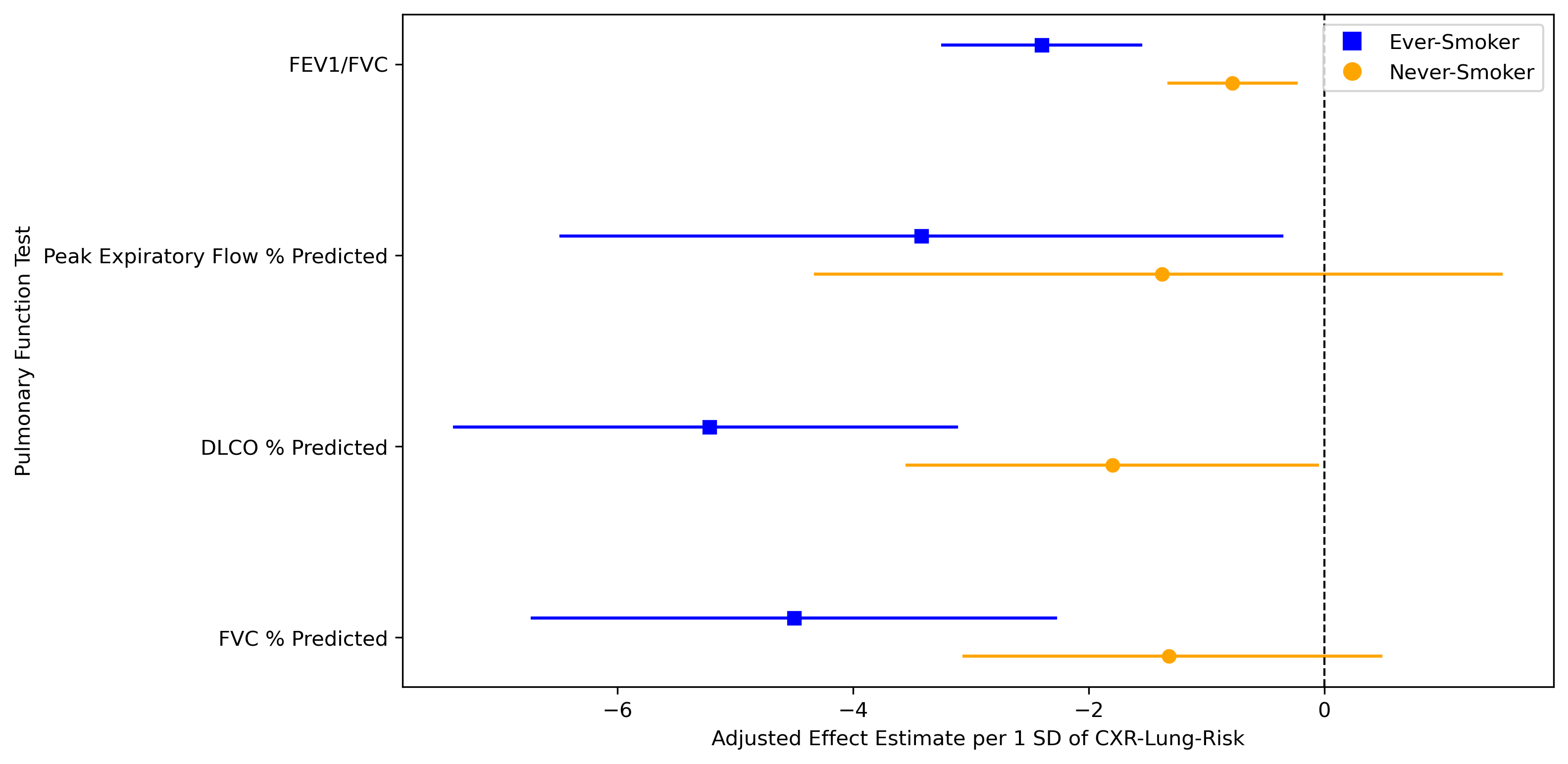

**Supplementary Figure 3. Association between pulmonary function tests and CXR-Lung-Risk in ever- (blue) and never- (orange) smokers in the Project Baseline Health Study controlled for lung disease**

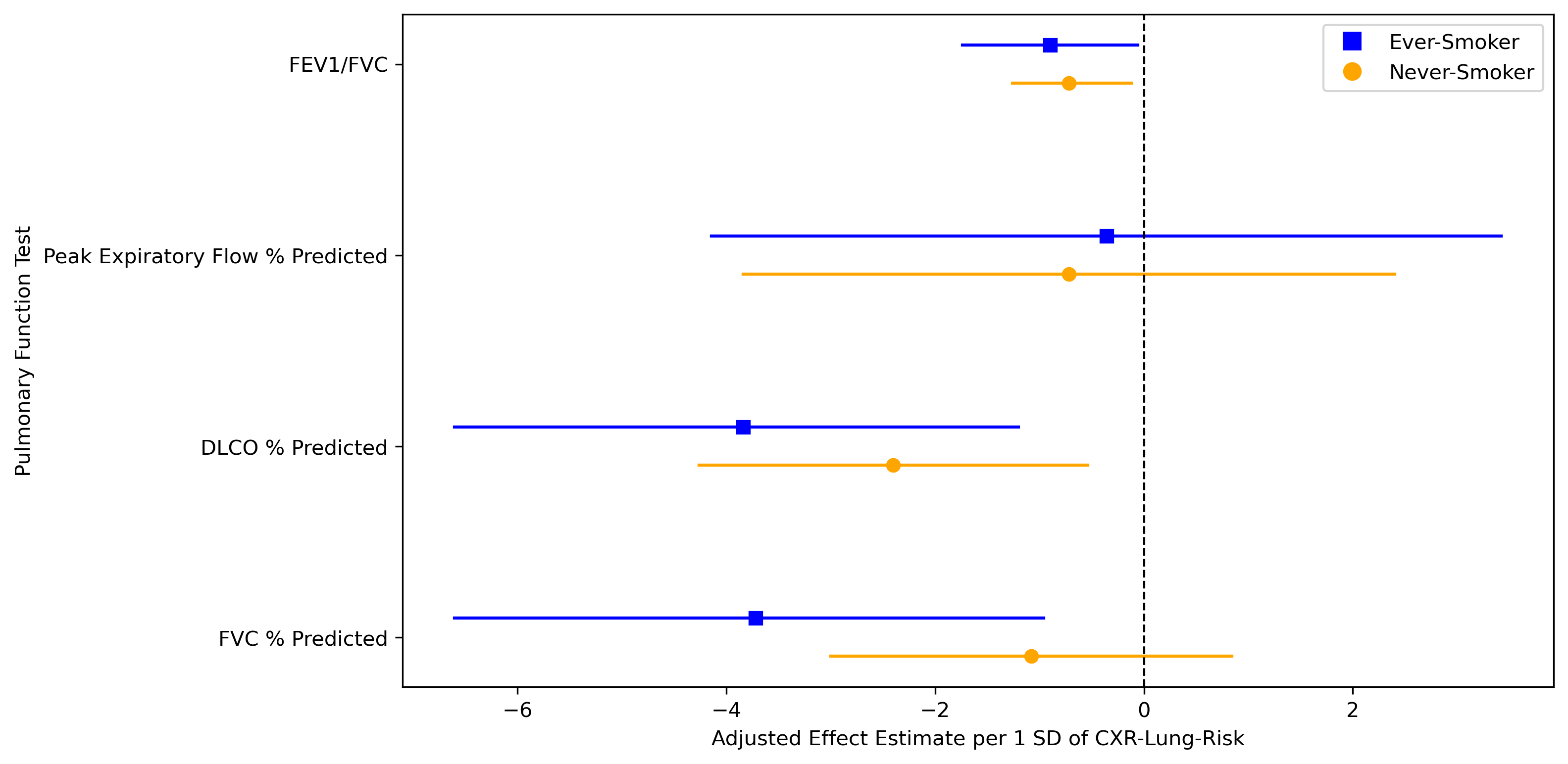

All estimates are adjusted for age, sex, body mass index (BMI), study site, frequency of smoking if the patient has smoked (everyday vs some days), and the presence of a lung disease (asthma, chronic obstructive pulmonary disease [COPD], chronic bronchitis, emphysema, pulmonary fibrosis, alpha-1 antitrypsin disease, sarcoidosis, and lung cancer).

**Supplementary Figure 4. Association of CXR-Lung-Risk with plasma protein concentrations (a) after adjustment for lung and kidney disease, (b) in ever-smokers, and (c) in never-smokers**

a

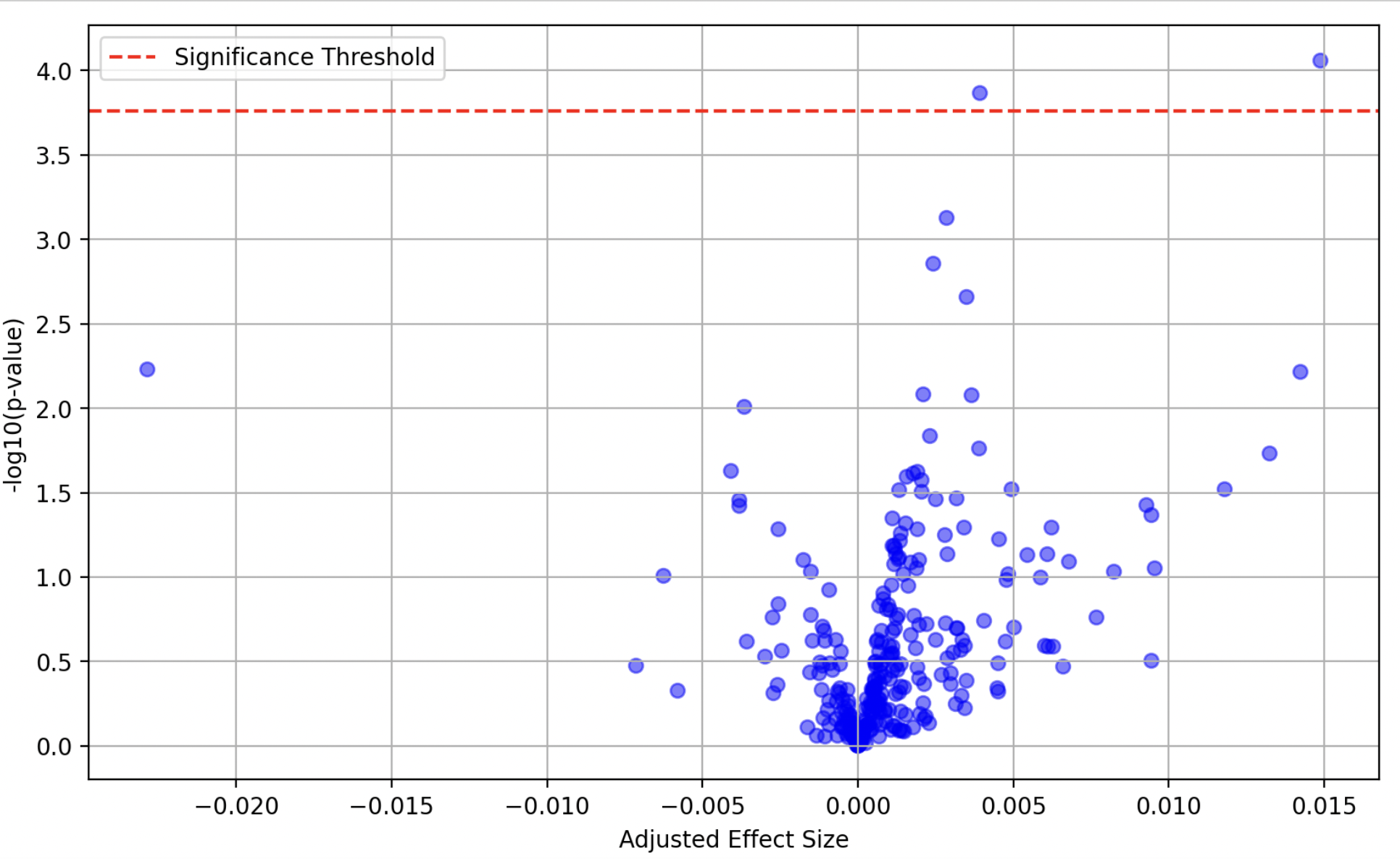

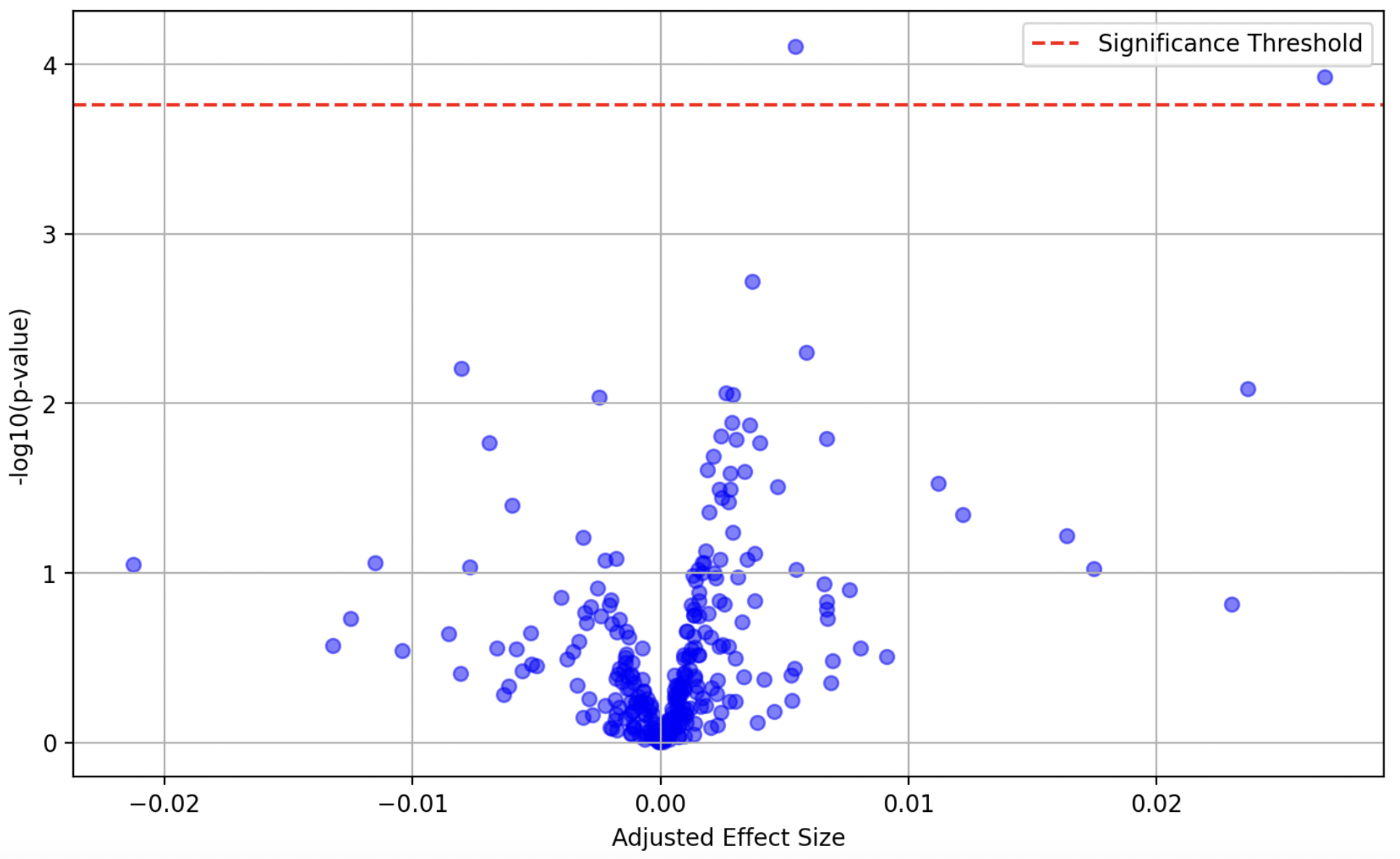

b

c

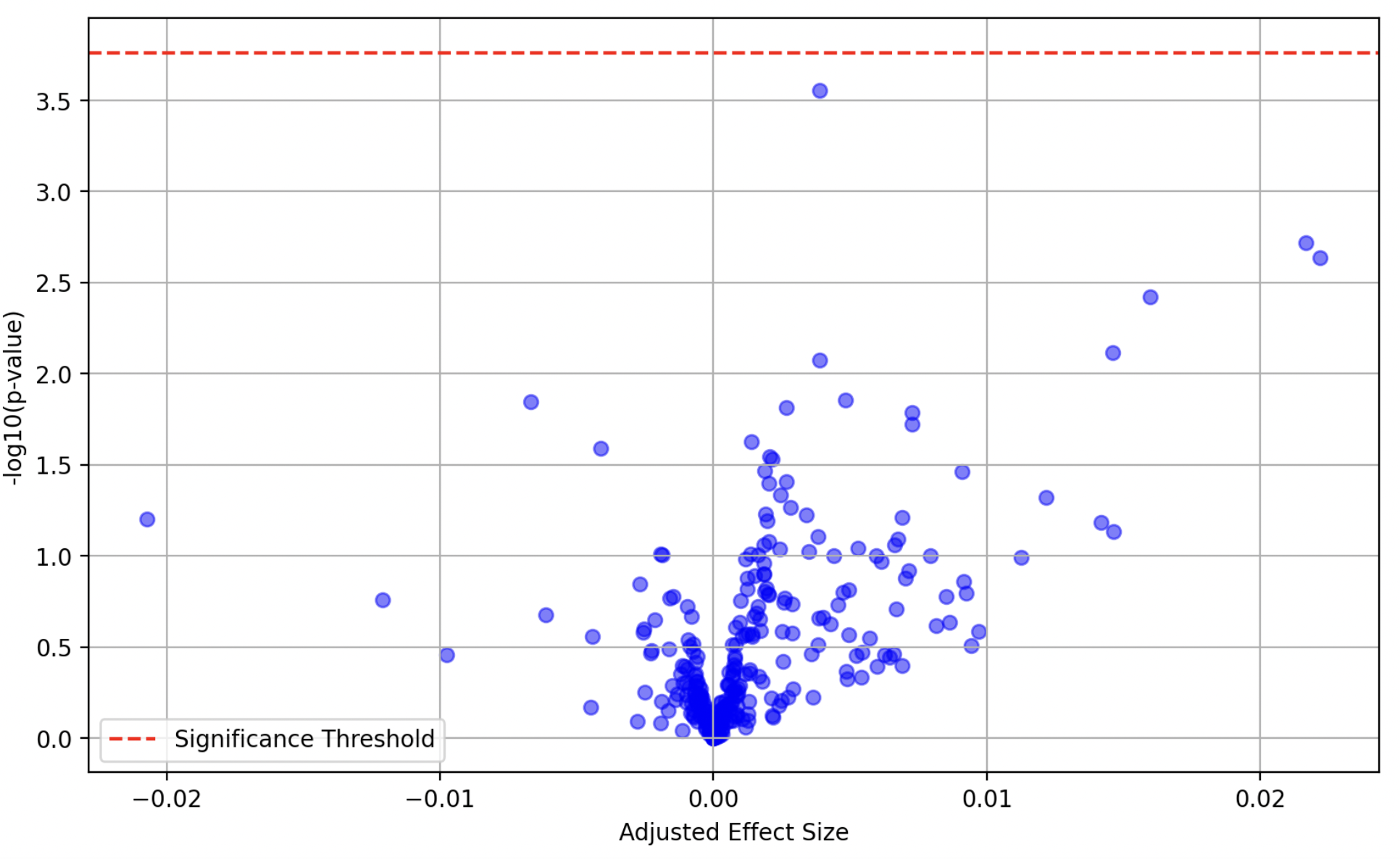
